# A personal network analysis of health-advice seeking and dietary influence in a rural community

**DOI:** 10.64898/2026.11.01.26366984

**Authors:** Marian-Gabriel Hâncean, Maria-Cristina Ghiţă, Matjaž Perc, Marius Geantă, Jürgen Lerner, Cosmina Cioroboiu

## Abstract

Health advice circulates through everyday relationships. The sources of informal health authority are less clear: the adviser’s knowledge and conduct, or their network position. We examine personal network data from 89 adults in a rural community (1,625 respondent-contact relationships). We distinguish contacts consulted for health advice from contacts who tried to change the respondent’s eating, model both relations with multilevel and penalised regressions, and test their differences. Both relations are embedded in frequent, close, and commensal ties and point to contacts more educated than the respondent. We find no characteristic that is more strongly associated with one relation than with the other. Health advice seeking also correlates with co-residence, the marital relationship, and responsibility for cooking. Respondents perceive their advisers as healthier, but advisers’ own reports give no evidence of healthier diets or lower body mass. Informal health authority here is constituted by relational position (intimacy, domestic food work, and a contact’s higher education relative to the respondent). We find no evidence that it rests on demonstrated conduct. Interventions that work through nominated advisers should not assume that being consulted implies expertise. Our design is cross-sectional and single-site. Future dyadic and longitudinal studies should test whether advice is followed.

## Introduction

Health-related practices are embedded in everyday social relationships. Individuals discuss health with relatives, friends, and other members of their personal networks, often before professional advice is sought (Freidson, 1960). Help-seeking is socially organised (McKinlay, 1973; Pescosolido, 1992), relationships are mobilised selectively for different purposes (Perry and Pescosolido, 2010), and health-advice relationships only partially overlap with close personal relationships (Reiner et al., 2025). Adviser selection depends on assessments of others’ knowledge and opportunities to access it (Borgatti and Cross, 2003). Understanding informal health advice therefore requires attention to the relationships through which particular people become sources of guidance, not only to individuals’ characteristics.

Unlike professional authority, grounded in credentials, informal advisory standing is established within relationships. It may reflect experiential knowledge of illness and healthcare (Prior, 2003), exemplary health conduct (Valente and Pumpuang, 2007), or relational position, such as being a spouse, cook, or educated relative. These possibilities matter: advisers chosen for their conduct may model healthier practices, whereas those chosen for their position need not. Distinguishing perceived conduct from reported conduct requires information from both the person seeking advice and the adviser. Overlapping personal network interviews can provide both.

Dietary practices are especially relevant because eating is organised through households, shared meals, and routines (Sobal et al., 2002). Food preparation is gendered caring labour (DeVault, 1991), and women often monitor family health (Gregory, 2005). Cooking may create opportunities for consultation without conferring control over consumption, challenging the notion of women as household food gatekeepers (McIntosh and Zey, 1989). Health-related social control also differs from support (Lewis and Butterfield, 2007): helping, persuading, and pressuring someone about food may have different consequences (Stephens et al., 2013).

These observations highlight a distinction that research on personal networks has paid little attention to: the difference between being consulted for health advice and trying to change another person’s eating. The first concerns the selection of a source of guidance. The second concerns a reported effort to modify behaviour. Both may occur within the same relationship, but they need not involve the same people, and both may be associated with gender, education, and relational proximity. Research on advice networks suggests that informal status, including relative expertise, affects advice relationships (Brooks et al., 2015). Educational differences within a relationship offer one observable dimension of such asymmetry, although education cannot be equated with health knowledge. Distinguishing advice from attempts at influence also matters for research on dietary behaviour in networks. Connected people often eat alike, but similarity can arise through influence, through choosing similar companions, or through shared circumstances (McPherson et al., 2001). Evidence from rural Eastern Europe documents such similarity in processed-food consumption within personal networks (Hâncean, Lerner et al., 2025). It shows that eating is socially patterned, but not who is influencing whom.

A rural community in Romania is an informative site for our questions. Rural populations in Eastern Europe are older than urban ones, carry a high burden of diet-related cardiovascular disease, and have more limited access to health services (Hâncean, McCarty et al., 2026). Informal advice is, therefore, likely to be a routine part of managing health. Households are frequently multigenerational, meals are shared, and residents know one another across overlapping ties. Personal network studies in this setting have documented the social clustering of smoking and diet (Mihaila et al., 2024; Hâncean, Lerner et al., 2025).

Our study asks three questions: (1) Which contact and relationship characteristics are associated with health advice seeking and attempted dietary influence? (2) How much do the two relations overlap, and do their associations differ in direct tests? (3) Are advisers healthier than those consulting them, according to respondents’ perceptions and advisers’ own reports? Addressing these questions, we make a threefold contribution. First, by examining whether advisory standing reflects perceived or self-reported health conduct or relational position. Second, by connecting gendered care and cooking responsibility to advice and attempted dietary influence. And, thirdly, by comparing these relations within the same personal networks. We underline that our article focuses on reported relationships and attempts, not on whether advice was followed or behaviour changed. Consequently, we cannot establish causality.

## Methods

### Study setting and design

This study uses personal network data collected in a rural commune of approximately 4,330 residents (3,808 adults) in Arge□ County, Romania, as part of an EU longitudinal project on primary cancer prevention (the 4P-CAN project; https://4p-can.eu/). The project follows an open-cohort longitudinal design with four waves of personal network interviews between September 2023 and September 2025 (Hâncean, McCarty et al., 2026). The analysis focuses on Wave 3 (March-April 2025), in which both health advice seeking and attempted dietary influence were recorded. Wave 4 (September 2025) examines the consistency of the advice findings under a different elicitation procedure and, for respondents interviewed in both waves, change over time.

Recruitment followed a link-tracing procedure from six purposively selected seeds who varied in sex, age, education, income, and employment. Participants recommended other eligible residents, whom the research team contacted. Eligibility required participants to be at least 18 years old, have lived in the community for at least six months, and provide informed consent. The sample is non-probability-based and not intended to represent the adult population. Wave 3 included 89 respondents, who described 1,625 respondent-alter relationships. Wave 4 included 87 respondents (67 returning, 20 first-time) and 2,529 relationships.

### Personal network data collection

Interviews were conducted face-to-face by trained interviewers using Network Canvas (Birkett et al., 2021), with name generators whose design evolved across waves (Hâncean, Lerner et al., 2026). Wave 3 asked for at least 25 contacts without a maximum cap. In Wave 3, respondents (egos) nominated adults (alters) living in the community with whom they communicated frequently, with separate prompts for household members, colleagues, relatives, and friends (with a distinction between contacts met face-to-face daily or weekly and those contacted daily or weekly by telephone or internet). Returning respondents generated their networks without being shown their earlier nominations. The median network contained 17 alters (range 2-48). In Wave 4, returning respondents reviewed their full historical roster. Namely, they reported contact frequency, emotional closeness, village residence, and years of acquaintance for each alter and answered the health-related items (shared meals, cardiovascular condition, and processed-food consumption) only for alters they met daily or weekly. First-time respondents followed the Wave 3 protocol, and only they were asked the dietary-influence item.

Individuals appearing across multiple networks or waves were tracked using harmonised identifiers matched manually from names, sociodemographic, and relational information. If an alter was nominated by two respondents, they contributed two observations while remaining identifiable as one person. For alters who also participated as respondents, interview data was linked to the reports about them, keeping perception and self-report distinct. Co-residence and kinship were taken as the last recorded value up to Wave 3 if not re-asked. The explicit kin relation was recorded only in Wave 1 for alters first nominated then. While households were not recorded as units, respondents reported whether they lived in the same house as each contact. Because individuals carried a single identifier across networks, we chained these reports: two respondents living with the same person were assigned to the same household, along with anyone else linked this way. Respondents with no co-residence links formed one-person households.

### Outcome measures

Two directed, respondent-reported relations were recorded for every alter in the Wave 3 roster. “Health advice seeking” indicates whether the respondent turned to this alter for health advice: a reported advisory relationship, not its frequency, content, or quality. “Attempted dietary influence” indicates whether this alter tried to change how the respondent ate: a reported attempt, regardless of whether the respondent changed their eating. Neither measure establishes the temporal sequence of the two activities.

### Explanatory variables

Alter characteristics, as reported by the respondent, were sex, age, education (up to eight years; nine years to post-secondary; tertiary), retirement, a cardiovascular condition, participation in sport, and whether the alter was also a Wave 3 respondent. Perceived health was measured by a ten-point rating of the alter’s health and by whether the alter was described as eating processed food daily or weekly. Relationship characteristics were co-residence, kinship outside the household, daily or weekly face-to-face contact, emotional closeness, sharing meals daily or weekly, and same sex. Wave 4 added years of acquaintance and village residence. Respondent characteristics were sex, age, education, household size, body mass index (from self-reported height and weight), and the number of eight processed-food categories consumed daily or weekly. Details on the study’s variables are available in the Supplementary Material (SM, Section 4.1).

#### Domestic roles

Among co-residents, we identified spouses and household cooks. Where available, Wave 2 supplied the cook’s person identifier. Otherwise, a respondent’s description (e.g., “my wife” or “my mother”) was matched to co-residents by relation, sex, and age. A unique match identified the cook, and ambiguous cases remained unassigned. Relative education classified alters as more, equally, or less educated than egos.

#### Self-reported differences

For 353 of the Wave 3 relationships, the contact had also been interviewed as a respondent, so both parties had reported their own processed-food consumption and body mass index. For each of these relationships, we subtracted the respondent’s value from the contact’s. A positive difference means that the contact reported eating more processed food or having a higher body mass index than the respondent. This allows advisers to be compared with the people who consult them.

### Statistical analysis

Figure 1 illustrates the data structure. The unit of analysis is the ego-alter tie: one observation per named contact, carrying the two outcomes and the characteristics of the contact and the relationship. Observations are dependent in two ways. Relationships reported by the same ego share that ego’s propensity to name advisers or influencers (Egos 2 and 3), and the same contact can appear in several ego-networks (Egos 7-8, 11-12, 15-16), including egos who are contacts, advisers, or influencers of other egos (Egos 5-6, 9-10, 13-14). The models address these dependencies with random intercepts or clustered standard errors for egos and alters, and the ego-to-ego ties with exponential random graph models.

**Figure 1.**
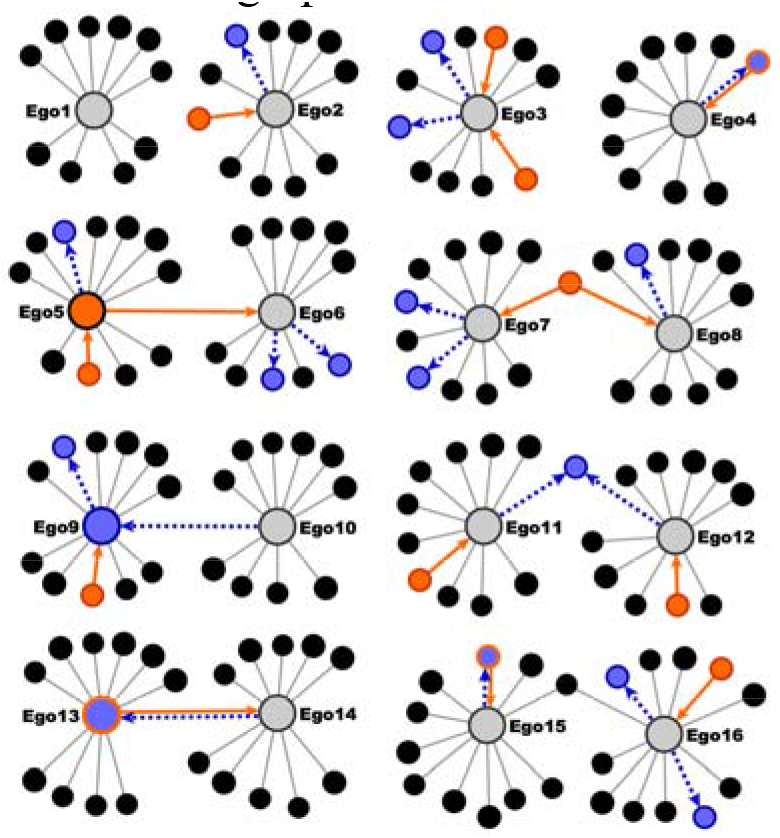
The data structure, illustrated with stylised examples. Large circles are egos (respondents), small circles are the alters they named, and grey lines are nominations. A blue dashed arrow runs from an ego to the alter it consults for health advice. An orange arrow runs from an alter who tried to change the ego’s eating to that ego. Blue marks advisers, orange marks sources of influence, and blue with an orange border marks both. Alters with neither status are black; egos with neither are grey. The same person may appear in several ego-networks, and egos may be advisers or influencers to one another.

#### Principal models

For health advice seeking, we fitted a logistic mixed-effects model (Model 1a, Table 2) with crossed random intercepts for respondents and for alters (Vacca et al., 2021). It estimates how the probability of being consulted depends on the alter’s characteristics, the relationship, and the respondent’s characteristics (the predictors are listed in Table 2). Model 1b replaces the alter’s education with its level relative to the respondent’s, asking whether people consult contacts more educated than themselves. Diagnostics are in the SM (Sections 15.1-15.5).

Attempted dietary influence was reported for only 78 relationships, too few for a reliable mixed model, so we used Firth’s penalised logistic regression (Model 2a), checked against ordinary logistic regression with respondent-clustered standard errors (Model 2b), and a mixed model with weak priors (SM, Sections 7 and 15.7). Model 2c is the relative-education version. Two further sets of models look inside the household. Among the 133 co-resident relationships, penalised regressions (Models 3a-3c, Table 3) tested whether the cook and the spouse are more likely to be consulted, or to attempt influence, than other household members, controlling for the alter’s sex and age. Among the 1,330 relationships with a recorded kin relation, mixed-effects models compared each type of relative with non-kin, before and after adding daily contact and closeness, to separate relatives consulted as everyday contacts from those consulted as kin (SM, Table S6).

Overlap between the two relations was described and then modelled (Model 2d, Table 2), with being an adviser as a predictor of attempted influence. Whether a characteristic matters more for one relation than for the other cannot be read off separately fitted models, because a difference between two estimates needs its own standard error. We therefore combined the two outcomes in a single “stacked” logistic regression (SM, Table S17): each relationship entered twice, once per outcome, and an outcome indicator was interacted with the 14 predictors of Model 2a so that the model estimates each predictor’s difference between outcomes directly, with standard errors clustered by respondent. The 14 differences in average marginal effects (influence minus advice), with delta-method confidence intervals and Holm-adjusted p-values, are the prespecified primary comparisons. As checks, we repeated the comparison on the log-odds scale, with two-way clustering by respondent and alter, and with a respondent-level bootstrap of 1,000 resamples (none failed). These tests compare the two outcomes under a common specification, not Model 1a with Model 2a.

Whether advisers are healthier than other contacts, or only perceived to be, was answered in three steps. First, two mixed models with the random intercepts of Model 1a tested whether respondents rate their advisers’ health higher, and describe them as eating processed food less often, than other contacts with the same characteristics. Second, in the 353 relationships where the contact had also been interviewed, regularised mixed models tested whether a contact who reports eating more processed food, or has a higher body mass index, than the respondent is more or less likely to be an adviser or an influencer, controlling for co-residence, shared meals, and closeness (SM, Table S7). Third, binomial mixed models with a household random intercept related the number of advisers, and of influencers, each respondent named, relative to network size, to the respondent’s own characteristics, including body mass index and processed-food consumption for influencers (SM, Table S8).

The 89 respondents also form a network among themselves (353 ties; 55 advice, 35 influence), which allows a test of whether advice is reciprocated or closes triangles, over and above individual and relational characteristics. Exponential random graph models were fitted to this directed network among respondents already in contact, with pairs not in contact fixed at zero, and terms for density, reciprocity, triadic closure, individual attributes, and co-residence. The reported advice model omits the closure term, which was not significant. Model terms, goodness of fit, and the influence model (which did not reproduce the out-degree distribution) are in the SM (Sections 12 and 15.9).

The advice models were re-estimated on Wave 4 data (SM, Table S2): Model 1a’s specification on the full rosters of the 67 returning respondents (2,115 relationships) and on the daily or weekly contacts of all 87 respondents (1,591, the only contacts with shared meals), and Model 1b’s on all 2,412 complete relationships. The influence item, asked only of the 20 first-time respondents (307 relationships, 25 nominations), was analysed with respondent-clustered logistic regression and a regularised mixed model including adviser status (SM, Table S9). For the 61 respondents in both waves, lagged regressions related Wave 4 processed-food count, body mass index, and dieting to their Wave 3 values and to having reported an influencer at Wave 3, with household-clustered standard errors. A simulation estimated power to detect reductions of 0.25 to 1 processed-food item (SM, Sections 14 and 15.10).

Results are reported as average marginal effects in percentage points with 95% confidence intervals: the change in the predicted probability for a 0 to 1 change in a binary predictor or a one-unit change in a continuous one, averaged over the observed relationships (from the fixed effects for mixed models). Odds ratios, random-effect estimates, and full diagnostics are in the SM. For replication, the data, code, and script are openly available (Hâncean, Ghita et al., 2026).

## Ethical approval statement

The study was approved by the Ethics Committee of the Center for Innovation in Medicine (EC-INOMED Decision No. D001/09-06-2023, No. D001/19-01-2024, and No. D002/03-02-2025).

## Results

### Descriptive statistics

Table 1 describes the 89 respondents (egos) and their 1,625 contacts (alters) in Wave 3. Egos turn to 163 alters (10.0%) for health advice; 73 egos (82%) name at least one adviser, typically one or two (Mdn = 1, range 0-7). Attempted dietary influence is rarer: 78 alters (4.8%), named by 38 egos (43%).

**Table 1.**
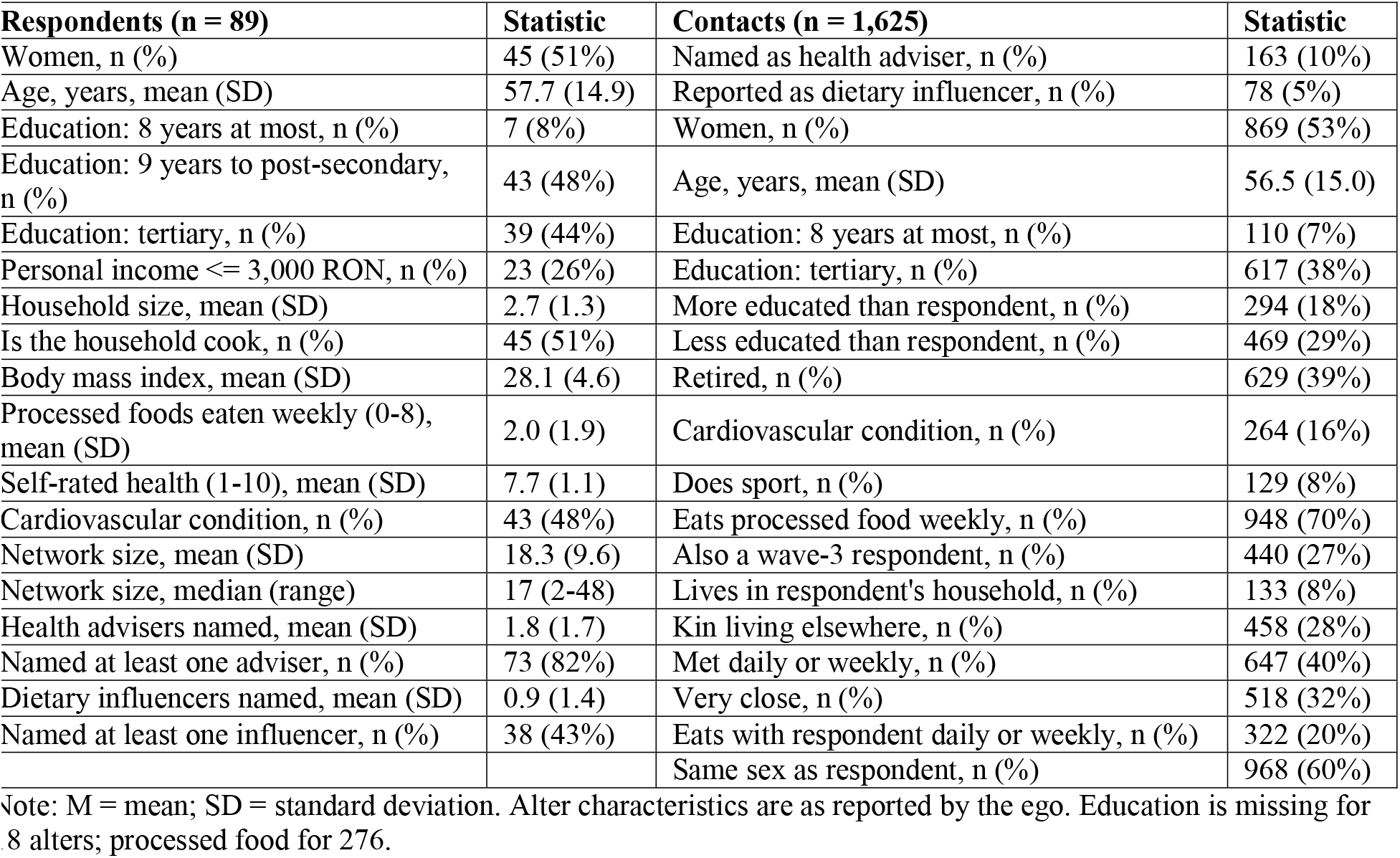
Descriptive statistics: respondents (egos) and their social contacts (alters), Wave 3.

| Respondents (n = 89) | Statistic | Contacts (n = 1,625) | Statistic |
|---|---|---|---|
| Women, n (%) | 45 (51%) | Named as health adviser, n (%) | 163 (10%) |
| Age, years, mean (SD) | 57.7 (14.9) | Reported as dietary influencer, n (%) | 78 (5%) |
| Education: 8 years at most, n (%) | 7 (8%) | Women, n (%) | 869 (53%) |
| Education: 9 years to post-secondary, n (%) | 43 (48%) | Age, years, mean (SD) | 56.5 (15.0) |
| Education: tertiary, n (%) | 39 (44%) | Education: 8 years at most, n (%) | 110 (7%) |
| Personal income <= 3,000 RON, n (%) | 23 (26%) | Education: tertiary, n (%) | 617 (38%) |
| Household size, mean (SD) | 2.7 (1.3) | More educated than respondent, n (%) | 294 (18%) |
| Is the household cook, n (%) | 45 (51%) | Less educated than respondent, n (%) | 469 (29%) |
| Body mass index, mean (SD) | 28.1 (4.6) | Retired, n (%) | 629 (39%) |
| Processed foods eaten weekly (0-8), mean (SD) | 2.0 (1.9) | Cardiovascular condition, n (%) | 264 (16%) |
| Self-rated health (1-10), mean (SD) | 7.7 (1.1) | Does sport, n (%) | 129 (8%) |
| Cardiovascular condition, n (%) | 43 (48%) | Eats processed food weekly, n (%) | 948 (70%) |
| Network size, mean (SD) | 18.3 (9.6) | Also a wave-3 respondent, n (%) | 440 (27%) |
| Network size, median (range) | 17 (2-48) | Lives in respondent's household, n (%) | 133 (8%) |
| Health advisers named, mean (SD) | 1.8 (1.7) | Kin living elsewhere, n (%) | 458 (28%) |
| Named at least one adviser, n (%) | 73 (82%) | Met daily or weekly, n (%) | 647 (40%) |
| Dietary influencers named, mean (SD) | 0.9 (1.4) | Very close, n (%) | 518 (32%) |
| Named at least one influencer, n (%) | 38 (43%) | Eats with respondent daily or weekly, n (%) | 322 (20%) |
|  |  | Same sex as respondent, n (%) | 968 (60%) |

Note: M = mean; SD = standard deviation. Alter characteristics are as reported by the ego. Education is missing for [illegible]8 alters; processed food for 276.

The two relations overlap but are not interchangeable. One in four of the contacts a respondent consults for health advice (41 of 163) has also tried to change the respondent’s eating, compared with one in forty of the contacts not consulted (37 of 1,462). Yet three in four advisers make no such attempt, and about half of all influence attempts come from people who are not consulted. In Model 2d (Table 2), being an adviser raises the probability of being reported as a dietary influencer by 7.4 percentage points (95% CI 2.0, 12.8), against a baseline of 4.8%.

**Table 2.** Models of health advice nominations and attempted dietary influence, Wave 3: average marginal effects in percentage points (95% CI)

|  | **Outcome: Health advice nominations** |  | **Outcome: Attempted dietary influence** |  |  |  |
|---|---|---|---|---|---|---|
|  | **Model 1a**<br>**All ties** | **Model 1b**<br>**Relative education** | **Model 2a**<br>**Firth** | **Model 2b**<br>**Respondent-clustered** | **Model 2c**<br>**Firth, relative education** | **Model 2d**<br>**Mixed, adviser included** |
| Contact is a woman | 7.5 (4.9, 10.2)*** | 7.1 (4.5, 9.7)*** | 2.5 (0.5, 4.4)* | 2.4 (0.6, 4.2)** | 1.9 (-0.0, 3.9) |  |
| Contact: tertiary education | 6.8 (3.7, 9.9)*** |  | 5.4 (3.1, 7.7)*** | 5.3 (2.2, 8.3)*** |  | 2.5 (0.7, 4.3)** |
| Contact: 8 years of school at most | -5.4 (-9.4, -1.5)** |  |  |  |  |  |
| Contact more educated than respondent |  | 5.5 (1.2, 9.7)* |  |  | 4.6 (1.3, 7.9)** |  |
| Contact less educated than respondent |  | -5.5 (-8.3, -2.7)*** |  |  | -1.5 (-3.8, 0.7) |  |
| Contact age (+10 years) | 1.2 (-0.3, 2.6) |  | -0.7 (-1.3, -0.0)* | -0.7 (-1.4, 0.0) |  | -0.3 (-0.7, 0.1) |
| Contact retired | -1.4 (-5.3, 2.5) |  |  |  |  |  |
| Contact: cardiovascular condition | 2.6 (-1.2, 6.4) |  | 2.9 (-0.7, 6.4) | 2.7 (-1.1, 6.6) |  |  |
| Contact does sport |  |  | 3.7 (-0.3, 7.8) | 3.5 (-0.7, 7.8) |  | 2.9 (0.1, 5.8)* |
| Contact is also a respondent | 2.2 (-0.8, 5.1) |  |  |  |  |  |
| Contact is a health adviser |  |  |  |  |  | 7.4 (2.0, 12.8)** |
| Same household (co-residence) | 12.8 (3.8, 21.7)** | 9.6 (2.1, 17.1)* | 4.2 (-0.9, 9.3) | 4.2 (-2.1, 10.4) | 2.5 (-1.6, 6.6) | 4.7 (0.6, 8.7)* |
| Kin, other household (living elsewhere) | 4.1 (0.4, 7.9)* | 3.6 (-0.0, 7.3) | -0.2 (-3.0, 2.6) | -0.2 (-3.4, 3.0) | -1.1 (-3.8, 1.5) |  |
| Daily or weekly contact | 6.3 (2.9, 9.6)*** | 7.1 (3.5, 10.6)*** | 3.5 (1.1, 5.8)** | 3.3 (-0.2, 6.9) | 3.6 (1.3, 5.9)** |  |
| Very close (emotional closeness) | 6.3 (2.3, 10.3)** | 6.7 (2.7, 10.7)** | 4.2 (1.3, 7.1)** | 4.0 (-0.4, 8.5) | 5.3 (2.4, 8.1)*** |  |
| Eats together daily or weekly (shared meals) | 8.3 (3.6, 13.1)*** | 8.7 (3.9, 13.6)*** | 3.4 (0.4, 6.4)* | 3.2 (-0.7, 7.2) | 4.5 (1.4, 7.7)** | 2.1 (0.1, 4.1)* |
| Same sex as respondent | 1.6 (-1.2, 4.4) |  | -0.3 (-2.4, 1.9) | -0.3 (-2.1, 1.6) |  |  |
| Respondent is a woman | -4.4 (-8.4, -0.4)* |  |  |  |  |  |
| Respondent age (+10 years) | 0.5 (-1.1, 2.0) |  | -0.6 (-1.3, 0.0) | -0.6 (-1.4, 0.2) |  | -0.5 (-1.2, 0.2) |
| Respondent: tertiary education | -3.1 (-7.0, 0.8) |  |  |  |  |  |
| Respondent household size (+1) | -2.3 (-3.7, -0.9)** |  | -0.3 (-1.2, 0.5) | -0.4 (-1.7, 1.0) |  |  |
| Respondent processed foods (+1 item) |  |  | 0.3 (-0.2, 0.9) | 0.3 (-0.5, 1.1) |  | 0.6 (-0.2, 1.4) |
| N ties / respondents | 1,606 / 89 | 1,607 / 89 | 1,607 / 89 | 1,607 / 89 | 1,607 / 89 | 1,607 / 89 |
| Advice nominations | 163 | 163 |  |  |  |  |
| Influence nominations |  |  | 78 | 78 | 78 | 78 |
| Random-effect SD: respondent / contact | 1.06 / 0.00 | 1.31 / 0.00 |  |  |  | 2.01 / 0.84 |
| Baseline probability | 10% | 10% | 4.8% | 4.8% | 4.8% | 4.8% |

Note. Average marginal effects: binary predictors 0 to 1, continuous predictors +1 unit; for Models 1a, 1b, and 2d from the fixed effects. Models 1a, 1b, and 2d: crossed random intercepts for respondents and for contacts. Models 2a and 2c: Firth’s penalised likelihood. Model 2b: maximum likelihood with standard errors clustered by respondent. Empty cells: predictor not in the model. Baseline: observed share of relationships with the outcome (see text). \* p < 0.05, \*\* p < 0.01, \*\*\* p < 0.001 (two-tailed).

### Who is consulted for health advice?

Effects should be read against the baselines of 10.0% (advice) and 4.8% (influence): an effect of +5 percentage points means a predicted probability 5 points higher for relationships with a characteristic than for otherwise similar relationships without it. Table 2 reports the models of both outcomes. Models 1a and 1b concern health advice. Female alters (Model 1a: AME = +7.5, 95% CI 4.9, 10.2) and alters with tertiary education (+6.8, 95% CI 3.7, 9.9) are more likely to be consulted, whereas alters with at most eight years of schooling are less likely (−5.4, 95% CI −9.4, −1.5).

Co-resident alters (+12.8, 95% CI 3.8, 21.7), kin living elsewhere (+4.1, 95% CI 0.4, 7.9, p < 0.05), alters met daily or weekly (+6.3, 95% CI 2.9, 9.6), and very close alters (+6.3, 95% CI 2.3, 10.3) are all more likely to be consulted, and eating together daily or weekly has an association of its own net of these (+8.3, 95% CI 3.6, 13.1). Advice is nevertheless not confined to the household: 64% of advisers live elsewhere. On the ego side, women (−4.4, 95% CI −8.4, −0.4, p < 0.05) and egos in larger households (−2.3 per additional member, 95% CI −3.7, −0.9) name fewer advisers per contact.

Replacing the alter’s education by its level relative to the ego’s (Model 1b, Table 2), alters more educated than the ego are more likely to be consulted (+5.5, 95% CI 1.2, 9.7, p < 0.05), and less educated alters are less likely (−5.5, 95% CI −8.3, −2.7), controlling for co-residence, kinship, contact, closeness, and shared meals.

### Who attempts to change the ego’s diet?

Models 2a-2d (Table 2) concern attempted dietary influence. The principal estimates are from Model 2a. Alters with tertiary education (+5.4, 95% CI 3.1, 7.7) and female alters (+2.5, 95% CI 0.5, 4.4, p < 0.05) are more often reported as influencers, as are alters more educated than the ego (Model 2c: +4.6, 95% CI 1.3, 7.9). Daily or weekly contact (+3.5, 95% CI 1.1, 5.8), closeness (+4.2, 95% CI 1.3, 7.1) and shared meals (+3.4, 95% CI 0.4, 6.4) are positively associated in the penalised model, although their intervals include zero under respondent-clustered inference. Co-residence and kinship outside the household are not clearly associated with attempted influence.

The stacked model (SM, Table S17) shows no statistically significant difference between the two outcomes after Holm adjustment for the 14 comparisons. The strongest unadjusted evidence concerns alter sex, whose association is 4.1 points smaller for influence than for advice (95% CI −7.1, −1.1; unadjusted p = 0.008; Holm-adjusted p = 0.11), with no corresponding difference on the log-odds scale (ratio of odds ratios 0.74, 95% CI 0.40, 1.35). The largest differences in absolute terms, for co-residence (−8.2, 95% CI −20.6, 4.3) and shared meals (−5.1, 95% CI −11.1, 0.8), are imprecise. Additionally, two-way clustering and bootstrapping yield the same conclusions.

### The household cook and the spouse

Within the 133 co-resident ties (Table 3), the cook is consulted in 30 of 37 cases (81%), against 26 of 96 other household members (27%). Net of alter sex, age, and spousal status, being the cook raises the probability of being the ego’s adviser by 40 percentage points (Model 3a: 95% CI 20, 60), and being the spouse by 46 points (95% CI 27, 64). Respondent-clustered estimates (Model 3b) are practically identical. All 13 spouses who cook are advisers, compared with eight of the 11 who do not. The corresponding estimates for attempted dietary influence (Model 3c) are positive but imprecise (cook: +14, 95% CI −7, 34; spouse: +15, 95% CI −6, 35).

**Table 3.**
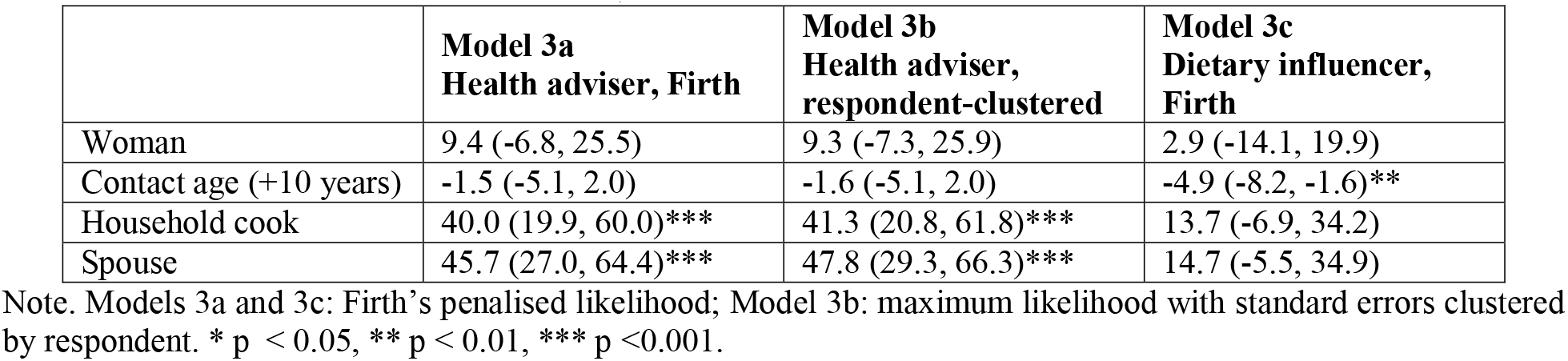
Within-household models among 133 co-resident alters (56 advice and 34 influence nominations): average marginal effects in percentage points (95% CI)

Among the 1,330 ties with a recorded kin relation, only the spouse has an association of its own: net of alter sex, education, age, contact, and closeness, a spouse is 46 points more likely than a non-kin alter to be consulted (95% CI 21, 72), whereas the associations for parents (+28 to +3) and children (+16 to +1) disappear once contact and closeness are controlled (SM, Table S6).

### Are advisers healthier than those who consult them?

Egos rate their advisers 0.22 points higher on a ten-point health scale (z = 2.5) and tend to describe them less often as eating processed food weekly (−9.5, 95% CI −20.0, 1.0; Wave 4: −10.7, 95% CI −20.9, −0.6; SM, Table S10). In the 353 ties in which both parties are respondents (SM, Table S7), the difference in self-reported processed-food consumption is not associated with being consulted (−0.7 per item, 95% CI −1.9, 0.5) and only weakly, in the expected direction, with attempted influence (−0.9, 95% CI −1.9, 0.1). The difference in BMI is associated with neither. With 55 and 35 nominations, these models are imprecise. At the ego level, the share of a network attempting dietary influence rises with the ego’s BMI (+0.3 points per unit, 95% CI 0.0, 0.5) and is unrelated to a cardiovascular condition (SM, Table S8).

Three further analyses are reported in full in the SM. Among respondents already in contact, advice ties concentrate within households (coefficient 2.94, SE 0.62) and show no evidence of reciprocity (−0.44, SE 0.82) or triadic closure (0.27, SE 0.38), although with 55 ties these tests have limited power (SM, Sections 12 and 15.9). In Wave 4, the same characteristics predict nominations, with larger associations for co-residence and closeness and a non-zero contact-level variance (SM, Table S2). Among the 20 first-time respondents, tertiary education (+12.5) and closeness (+12.0) again predict attempted influence (SM, Table S9). Among the 61 egos interviewed in both waves, those who reported an influencer in Wave 3 (n = 24) did not differ from the others in the change in processed-food consumption, BMI, or dieting by Wave 4. However, the design has a power of only 0.21 to detect a reduction of half a processed-food item and 0.61 for one item (SM, Sections 14 and 15.10).

## Discussion

Three findings stand out. Health advice seeking and attempted dietary influence were associated with the same social and relational characteristics (education, frequent interaction, closeness, shared meals), and advice seeking was also associated with co-residence, marital relationship, and responsibility for cooking. Although the two activities differed in prevalence and only partially overlapped, no characteristic was more strongly associated with one than the other. Respondents perceived their advisers as healthier, but the advisers’ own reports gave no evidence that they were. Informal health authority thus appears to rest on relational position and perceived characteristics rather than on demonstrably healthier conduct.

### The relational organisation of health advice

Most respondents named an adviser, yet only one in ten ties involved advice. This selective mobilisation is consistent with work on functional specificity in personal networks (McKinlay, 1973; Pescosolido, 1992; Perry and Pescosolido, 2010; Reiner et al., 2025).

Two features of the selection deserve emphasis. First, respondents were more likely to consult contacts more educated than themselves and less likely to consult those less educated, controlling for co-residence, kinship, contact, closeness, and shared meals. This pattern also held in Wave 4. This is compatible with the idea that advice seeking involves assessments of relative competence (Agneessens and Wittek, 2012; Blau, 1954), although educational differences may also carry social hierarchies, confidence in communication, or the perceived legitimacy of offering guidance rather than health expertise. Second, shared meals showed an association of their own after controlling for co-residence, contact frequency, and closeness. Also, in Wave 4, commensality was the strongest relationship feature among daily contacts. Meals create repeated occasions on which eating is visible and can potentially affect health (Sobal and Nelson, 2003), although we cannot establish the direction of the association here. Health advice was not confined to the household: nearly two-thirds of advisers lived elsewhere, and relatives living elsewhere were consulted more often than non-kin.

### Gendered care, domestic work, and advisory standing

Female contacts were more likely to be nominated as advisers, controlling for education and relational characteristics. This is consistent with the gendered distribution of domestic care, food provision, and responsibility for family health (Umberson, 1992; Clark, 1995). Similar results were also reported in other cultural settings (Reiner et al., 2025). The household analyses provide more direct evidence. Among co-resident contacts, the cook and the spouse were much more likely to be nominated, controlling for the alter’s sex and age. The kin analysis reinforces the point: the spouse retained a substantial association after controlling for contact and closeness, whereas parents and children were consulted only because they were everyday, close contacts.

Our findings do not suggest that food preparation confers authority over dietary decisions. The distinction between performing food-related care and controlling consumption is supported in the literature (Blissett and Haycraft, 2008). Cooking responsibility and spousal status were strongly associated with being consulted, whereas their associations with attempted dietary influence were positive but too imprecise to characterise. Whether domestic roles also carry attempts to regulate another person’s eating remains open.

### Advice seeking and attempted dietary influence: distinct activities, similar relationships

The two relations differed in prevalence and only partially overlapped. Yet the overlap was far greater than their prevalence alone would produce. Being an adviser raised the probability of being reported as an influencer from under five per cent to about twelve per cent, controlling for other characteristics. Some relationships thus accommodate several forms of health-related involvement, even though someone may not request or welcome an attempt to change their eating.

None of the 14 predictor-specific differences was statistically significant after Holm adjustment. The two effects that were suggestive before adjustment (the alter’s participation in sport, associated with influence but not advice, and the respondent’s household size) were not significant after it. The findings therefore do not establish that the two activities have different relational foundations, nor does the absence of significant differences demonstrate equivalence. Among the measured characteristics, both activities were associated with the same social and relational contexts. The same relationships create opportunities for both guidance and attempts at change. Whether an interaction is experienced as advice, care, persuasion, or pressure may depend on unmeasured content.

One asymmetry appeared on the respondent side. The share of a network reported attempting dietary influence rose with the respondent’s body mass index and tended to rise with processed-food consumption, but it was unrelated to a cardiovascular condition. Among first-time respondents in Wave 4, body mass index was again associated with being an influence target. Research on health-related social control has found that attempts to regulate a partner’s behaviour are directed at those perceived to need it and are shaped by the target’s weight (Novak and Webster, 2011; Brunson et al., 2014). Our results suggest that dietary pressure in this community responds to visible body size rather than diagnosed risk, although the estimates are based on 89 respondents.

### Perceived health, self-reported behaviour, and the basis of informal authority

Respondents rated nominated advisers as healthier than other contacts and tended to describe them as eating processed food less often. This is consistent with the possibility that perceived conduct contributes to advisory standing (Valente and Pumpuang, 2007). The analyses using independently reported characteristics did not provide corresponding evidence. Namely, among relationships in which both parties were interviewed, differences in self-reported processed-food consumption and body mass index were not associated with being consulted, and only weakly with attempted influence. This should be interpreted with care, as self-reports were available only when both individuals participated, nomination counts were small, and the two measures only partially capture health-related knowledge or exemplary conduct. The absence of an association is a lack of evidence that advisers differ, not evidence that they are alike. A US survey found that people who give health advice to many others report somewhat healthier behaviours (Colón-Ramos et al., 2009). Our comparison, using the reports of both parties to the same relationship, found no such difference.

Professional authority is institutionally credentialled, whereas informal advisory standing must be established within relationships (Freidson, 1960) and may reflect perceived knowledge, experience of illness, educational position, trust, familiarity, or responsibility for care (Prior, 2003). Our findings do not isolate these sources, but they show that the three positional ones (i.e., relative education, relational proximity, and domestic responsibility) are each associated with being consulted, while the exemplary one (healthier conduct) is perceived and not confirmed. Informal health authority in this community is thus not an attribute of the adviser but a feature of where the adviser stands in relation to the person asking. Identifying frequently consulted individuals reveals socially recognised sources of guidance, not people who hold accurate information or follow healthier practices. Interventions working through personal networks therefore need to account for what advisers communicate and not assume that standing indicates expertise.

### Network structure, consistency across waves, and change over time

Advice ties among respondents clustered within households but showed no clear reciprocity or triadic closure. Nevertheless, the small network limits inference. Wave 4 showed broadly similar correlates, but its changed roster and overlapping respondents make it a within-sample consistency check, not an independent replication. The 61-person panel found no association between reported influence and later dietary or BMI outcomes. On the other hand, this analysis had limited power and recorded attempts rather than their intensity or reception. Consequently, it does not establish that influence is ineffective.

### Limitations

The respondents were recruited by link-tracing in one rural community, not by probability sampling. The main findings come from comparisons among the contacts within each respondent’s network. Thus, the two outcomes and the contact characteristics were recorded for every contact named. The contacts are those the respondents named under the generator’s rules, so they represent frequent local relationships rather than all of a person’s ties. The outcomes are what respondents reported, not observed interactions, and the characteristics they reported about their contacts may be biased by the relationship itself (e.g., advisers may be described more favourably). The cross-sectional design cannot establish causal direction. Attempted dietary influence was reported for only 78 relationships. Penalised estimation addressed this, but several estimates for this outcome remained imprecise. Some analyses are based on subsets or reconstructed measures, i.e., the cook was identified for 38 respondents, kin relations only for contacts first named in Wave 1, and self-reports only for the 353 relationships in which both parties were interviewed. Future research could interview both parties in each relationship, ask respondents directly how knowledgeable and trustworthy they consider each contact, and follow networks over longer periods.

## Conclusion

Health advice seeking and attempted dietary influence differ in prevalence and overlap only partially. Yet they share the same social and relational correlates. Health advice seeking is connected to relative educational standing, relational proximity, and domestic roles, including the marital relationship and responsibility for cooking. Attempts at dietary influence seem to be directed at heavier respondents (higher BMI) and not at those with diagnosed conditions. Respondents perceive advisers as healthier, but advisers’ own reports do not confirm this. Informal health authority in this community rests on where people stand in relation to one another more than on their observable health-related conduct.

## Data Availability

The data and code supporting the findings of this study are openly available on Zenodo at https://doi.org/10.5281/zenodo.23807481.

## Ethics approval statement

All research procedures complied with the Declaration of Helsinki and the European General Data Protection Regulation (GDPR). The study was approved by the Ethics Committee of the Center for Innovation in Medicine (EC-INOMED Decision No. D001/09–06–2023, No. D001/19–01–2024, and No. D002/03–02–2025).

## CRediT authorship contribution statement. M.-G.H

Writing-review & editing, Writing-original draft, Visualization, Methodology, Investigation, Formal analysis, Data curation, Conceptualization. **M.-C.G**.: Writing-review & editing, Writing-original draft, Formal analysis, Data curation, Conceptualization. **M.P**.: Writing-review & editing, Writing-original draft, Validation, Methodology, Conceptualization. **M.G**.: Writing-review & editing, Writing-original draft, Validation, Project administration, Funding acquisition, Conceptualization. **J.L**.: Writing-review & editing, Writing-original draft, Validation, Methodology, Conceptualization. C.C.: Writing-review & editing, Writing-original draft, Resources, Project administration.

## Data availability

Data, code, and supplementary material are openly available as Hâncean, M.-G., Ghita, M.-C., Perc, M., Lerner, J. and Geanta, M. (2026) Data for a personal network analysis of health-advice seeking and dietary influence in a rural community. Zenodo, [Dataset]. https://doi.org/10.5281/zenodo.23807481

